# A single-centre, observational study to evaluate immune response to Covid-19 vaccines in immunocompromised patients with haematological disorders (COVAC-IC)

**DOI:** 10.1101/2022.11.16.22282121

**Authors:** COVAC-IC investigators, Deepak Chandra, Lucy O’ Mara, Lucy Bailey, Mathew Aspey, Md Asaduzzaman, Krishna Banavathi, Simon Lea, Rob Bowler, Jayasekara Prasangika, Aviva Ogbolosingha, Sarah Goddard, Neil Phillips, Fauzia Wasim, Buddhika Badugama, Nausheen Kamran, Kumari Perera, Fehmida Bano, Srinivas Pillai, Peter Dyer, Muzna Aquil, Alda Remegoso, Judith Lee, Keira Watts, Kamaraj Karunanithi

## Abstract

**Objective:** To evaluate immunological response to Covid-19 vaccines in immunocompromised haematology patients and compare with immunocompetent healthy controls

**Design:** We compared total Anti-SARS-CoV-2 spike antibody and T cell response in 45 immunocompromised haematology patients with 30 healthy adults following 2 doses of Covid-19 vaccine for 3 -5 months at 30 day intervals

**Setting:** Single Centre, University Hospital, United Kingdom, March 2021-December 2021

**Main Outcome measures:** Peak quantitative total spike-specific antibody and cellular responses

**Results:** We found

1. Non - significant difference in T cell and total Anti-SARS-CoV-2 S antibody response between study and control group patients
2. Six (13%) study group participants did not have detectable Total Anti-SARS –Cov-2 S antibodies at any time point throughout the study monitoring period.
3. Three (7%) of the study group participants had no response, even after additional booster doses of Covid-19 vaccine.
4. All (100%) of the control group had detectable Anti-SARS-Cov-2 S antibodies after 2 doses of Covid-19 vaccine.
5. No participant died or was hospitalised due to severe Covid-19 infection during the study period. This included study group participants who had no antibody response at any time point.

**Conclusions:** Though there was a non - significant difference in T cell and total Anti-SARS-CoV-2 S antibody response between immunocompromised patients and healthy controls this did not result in any severe infection or Covid-19 related mortality in our study cohort. We did not identify any patient-specific factor (age, gender), specific haematological condition or treatment as determinant of response. Covid-19 vaccination was well tolerated without major side effects in both groups.

**What was already known about this topic:** prior to starting this study there were no studies to confirm immunological response following Covid-19 vaccination in immunocompromised haematology patients. During the conduct of our study there have been publications from researchers confirming blunted serological response in 62-66% of immunocompromised haematology patients compared to 74-95% in healthy controls.

**What this study adds:** Our study did not identify a significant difference in serological or T cell response between immunocompromised and healthy groups. Though 13% of immunocompromised patients had no response to Covid-19 vaccination none of them suffered from severe Covid-19 infection. We believe T cell response to Covid-19 vaccination has an important role in providing protective efficacy against Covid-19.

## Introduction

Severe Acute Respiratory Syndrome coronavirus 2 (SARS-Cov-2; Covid-19) infection and disease has affected millions of persons worldwide. The World Health Organisation declared it as a global pandemic in March 2020. Since the beginning of the pandemic over 22million people have been infected with Covid-19 in the United Kingdom with over 175,000 deaths (until June 2022) [1].

Patients with haematological diseases often have impaired immunity due to their blood disorder and/or as a consequence of treatment with chemotherapy or immunotherapy. Patients remain immunocompromised for several months after diagnosis and treatment.

Covid-19 infection in immunocompromised patients was reported early in the pandemic to be more severe with higher mortality [2]. These individuals were a high risk group to be prioritised for vaccination against Covid-19. Vaccines against Covid-19 were developed and introduced with great speed to control the pandemic. When introduced, these vaccines had not been tested in immunocompromised individuals. Their protective efficacy, safety and durability in immunocompromised individuals were therefore unknown.

## Methods

We initiated this study in March 2021 to compare immunological response to Covid-19 vaccine in immunocompromised patients with haematological disorders and compared it with healthy controls (COVAC-IC, ClinicalTrials.gov Identifier: NCT04805216). The study was conducted in accordance to declaration of Helsinki and good clinical practice following REC approval (London Bridge Research Ethics Committee, reference 21/HRA/0304). Patients with haematological disorders and clinically assessed to be immunocompromised either due to their haematological condition or treatment constituted the study group. Healthy volunteers assessed to have normal immunity were recruited as the control group. Between 3 and a maximum of 5 venous blood samples were obtained from all participants at 30 day intervals following their 2nd dose of Covid-19 vaccination. Due to changes in public health vaccination schedule, participant entry criteria and blood sample scheduling had to be modified (with REC and sponsor approval). Study modification allowed recruitment of participants at the nearest 30 day time–point after the 2nd dose of Covid-19 vaccination. Immune response was monitored for 3-5 months at 30 day time intervals. The blood samples were tested for Total Anti-SARS-CoV-2 to S and N-antigen (Roche Elecsys®) and for SARS-CoV-2 S1/S2 IgG antibody (DiaSorin Liaison®). Seropositivity was defined as SARS-CoV-2 spike-receptor binding domain specific total IgG antibody above the threshold of detection for the assay. T-cell response was assessed by Interferon gamma release assay (IGRA) using the Qiagen QuantiFERON SARS-Cov-2 RUO® blood collection tubes and ELISA. A positive response was defined as increase in frequency of SARS-CoV-2 specific CD4/CD8 T cells after vaccination. We had initially planned to perform the T-cell analysis at 3 time points however lack of kit availability only allowed us to do this test on 1st blood sample (participant entry to the study). Statistical techniques including data summarisation, graphs and hypothesis testing were performed to make valid conclusions.

## Results

Forty-five patients (25 male, 20 female) age range 19-78 years (median 67 years, IQR 13 years) with impaired immunity due to haematological disorder or treatment and 30 immunocompetent healthy individuals (6 male, 24 female) age range 18-74 years (median 50 years, IQR 23 years) were recruited to the study between April – December 2021 (Table 1). All participants had received at least 2 doses of Covid-19 vaccine (Either ChAdOx1-S (AstraZeneca) or BNT162b2 mRNA vaccine (Pfizer)) before entry into the study. Three hundred and thirty two blood samples were collected and analysed to evaluate immune response as per the study protocol. Comparison was made of Total Anti-SARS-CoV-2S antibody between study and control groups at specific time points after 2^nd^ dose of Covid-19 vaccination. T cell response was assessed on the first blood test at entry to the study.

**Table 1:** Details of patient cohort

|  | Patient cohort (study group) | Control group |
| --- | --- | --- |
| Age |  |  |
| Median | 67 | 49.5 |
| IQR | 13 | 22.5 |
| Range | 19-78 years | 18-74 years |
| Sex |  |  |
| Male | 25 (56%) | 6 (20%) |
| Female | 20 (44%) | 24 (80%) |
| Vaccine received |  |  |
| Pfizer | 14 (31%) | 14 (47%) |
| Astra Zeneca | 31 (69%) | 16 (53%) |
| **Time (days) after 2<sup>nd</sup> dose vaccine to 1<sup>st</sup> blood test** | **Number of patients (study group)** | **Number of patients (control group)** |
| 15 | 7 | 7 |
| 30 | 13 | 8 |
| 150 | 4 | 3 |
| 180 | 10 | 5 |
| 210 | 4 | 4 |
| 240 | 5 | 2 |
| 270 | 1 |  |
| 330 | 1 | 1 |
| **Underlying haematological diagnosis** | **Study group** | **Control group** |
| Acute Leukaemia | 3 (7%) | Healthy volunteers |
| Chronic Lymphocytic Leukaemia | 11 (24%) |  |
| Hodgkin's Lymphoma | 2 (4%) |  |
| Non-Hodgkin's Lymphoma | 5 (11%) |  |
| Multiple Myeloma | 24 (53%) |  |
| Non-Malignant Immunosuppressant Therapy | 3 (7%) |  |
| Other | 15 (33%) |  |

Six (13%) study group participants did not have detectable Total Anti-SARS –Cov-2 S antibodies at any time point throughout the study monitoring period (Table 2). Three (7%) of the study group participants had no response, even after additional booster doses of Covid-19 vaccine. All (100%) of the control group had detectable Anti-SARS-Cov-2 S antibodies after 2 doses of Covid-19 vaccine.

**Table 2:**
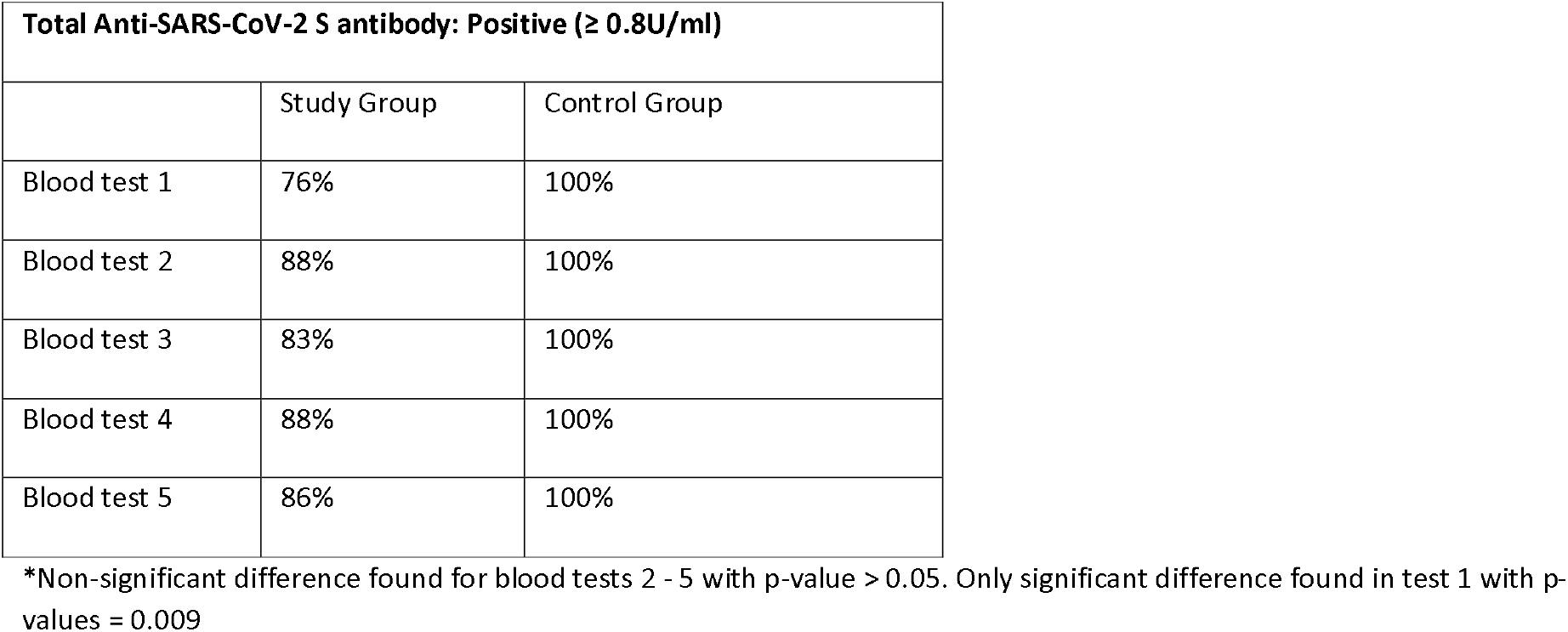
Total Anti-SARS-Cov-2 S antibody positive response

The T cell response assessed by IGRA was reactive in 53% of study group patients compared to 77% of the control group at least 30 days after the 2^nd^ Covid-19 vaccine. This difference was not significant (p-value = 0.098 > 0.05) (Table 3).

**Table 3:** T cell response assessed by IGRA assay

| T cell response QuantiFERON SARS-Cov-2: Reactive (%) |  |  |
| --- | --- | --- |
|  | Study Group | Control Group |
| Blood test 1 | 53% (n=30) | 77% (n=27) |
| (at least 30days after 2<sup>nd</sup> dose of Covid-19 vaccine) |  | 78% (n=27) |

\*Non-significant difference found with p-value = 0.098.

We found non-significant difference in T cell and total Anti-SARS-CoV-2 S antibody response between study and control group patients (p-value > 0.05 other than for test 1 where p-value < 0.05) (Table 2 & Table 3).

Different vaccination times, doses, timing of blood test and natural infection with Covid-19 were variables which would have affected the vaccine response. Other factors include age, haematological disorder, immunosuppressive treatment (and its timing).

When the study subgroup that had not received additional doses of Covid vaccine or had suffered from natural infection was compared with the control group the difference was still not significant (p-value > 0.05).

We tried to identify clinical characteristics which were determinants of response by comparing study subgroups with poor or good antibody response. Poor responders were defined as individuals who had anti-SARS CoV-2 S antibodies below the level of detection at all-time points. Good responders were responders with antibody levels above the upper limit of detection at all-time points. Neither a specific haematological diagnosis nor chemotherapy or immunotherapy was identified as a determinant of response (Table 4 and Table 5).

**Table 4:** Study group (immunocompromised) patients with poor antibody response to Covid-19 vaccination

| Study ID# | Age range (yr)/Sex | Clinical (diagnosis; treatment) | Total Antibody to Spike protein (result U/ml; days after 2nd dose of covid vaccine) |  |  |  |  |
| --- | --- | --- | --- | --- | --- | --- | --- |
|  |  |  | BT1 | BT2 | BT3 | BT4 | BT5 |
| P012 | 71-75/F | Stage B progressive CLL; Ibrutinib | <0.4; 30d | <0.4; 64days after 2<sup>nd</sup> dose | <0.4; 90days after 2<sup>nd</sup> dose | <0.4; 120days after 2<sup>nd</sup> dose | ND |
| P014 | 71-75/F | Acute Myeloid Leukaemia; Azacitidine, venetoclax | <0.4; 30days | <0.4; 60days after 2<sup>nd</sup> dose | <0.4; 89days after 2<sup>nd</sup> dose | <0.4; 118days after 2<sup>nd</sup> dose | ND |
| P018 | 71-75/F | CLL, chronic ITP; low dose pred, MMF | <0.4; 34days after 2<sup>nd</sup> dose | <0.4; 58days after 2<sup>nd</sup> dose | <0.4; 90days after 2<sup>nd</sup> dose | <0.4; 119days after 2<sup>nd</sup> dose | ND |
| P065* | 76-80/M | High risk Burkitts Lymphoma; RCHOP, EPOCH-R | <0.4; 210days after 2<sup>nd</sup> dose, 20days after 3<sup>rd</sup> dose | <0.4; 240days after 2<sup>nd</sup> dose, 50days after 3<sup>rd</sup> dose | <0.4; 270days after 2<sup>nd</sup> dose, 80days after 3<sup>rd</sup> dose | <0.4; 300days after 2<sup>nd</sup> dose, 110days after 3<sup>rd</sup> dose | <0.4; 330days after 2<sup>nd</sup> dose, 140days after 3<sup>rd</sup> dose, 20days after 4<sup>th</sup> dose |
| P074* | 66-70/M | CLL; No treatment | <0.4; 240days after 2<sup>nd</sup>, 50days after 3<sup>rd</sup> dose | <0.4; 270days after 2<sup>nd</sup>, 80days after 3<sup>rd</sup> dose | <0.4; 300days after 2<sup>nd</sup>, 110days after 3<sup>rd</sup> dose | <0.4; 330days after 2<sup>nd</sup>, 140days after 3<sup>rd</sup> and 12 days after 4<sup>th</sup> dose | <0.4; 360days after 2<sup>nd</sup>, 170days after 3<sup>rd</sup> and 42days after 4<sup>th</sup> dose |
| P076* | 61-65/M | CLL; Acalabrutinib | <0.4; 240days after 2<sup>nd</sup>, 40days after 3<sup>rd</sup> dose | <0.4; 270days after 2<sup>nd</sup>, 70days after 3<sup>rd</sup> dose | <0.4; 300days after 2<sup>nd</sup>, 100days after 3<sup>rd</sup>, 5days after 4<sup>th</sup> dose | <0.4; 330days after 2<sup>nd</sup>, 130days after 3<sup>rd</sup>, 35days after 4<sup>th</sup> dose |  |
| *no response despite 4 doses of covid vaccination |  |  |  |  |  |  |  |
| #study ID's are not known to anyone outside the research group |  |  |  |  |  |  |  |

**Table 5:**
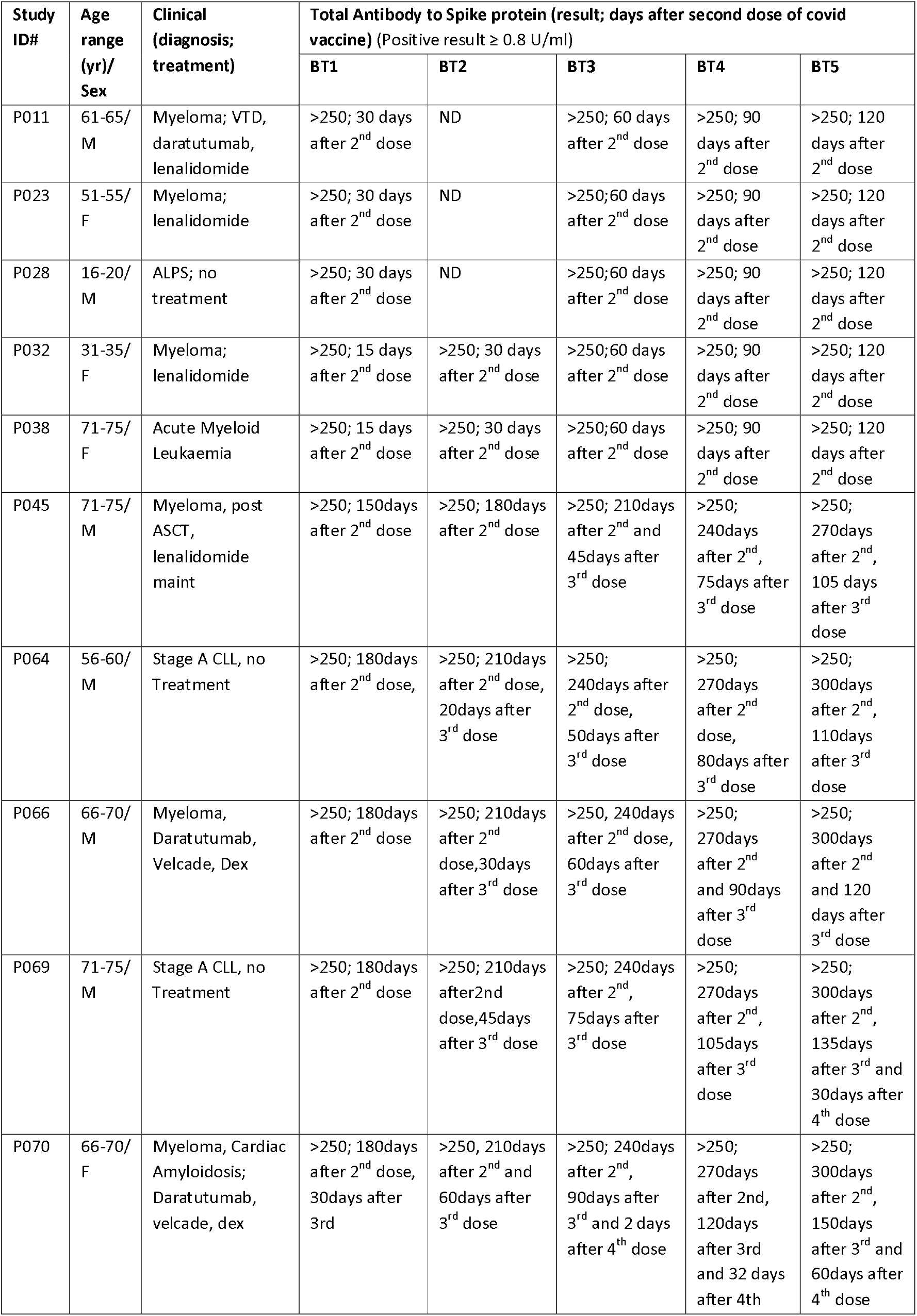
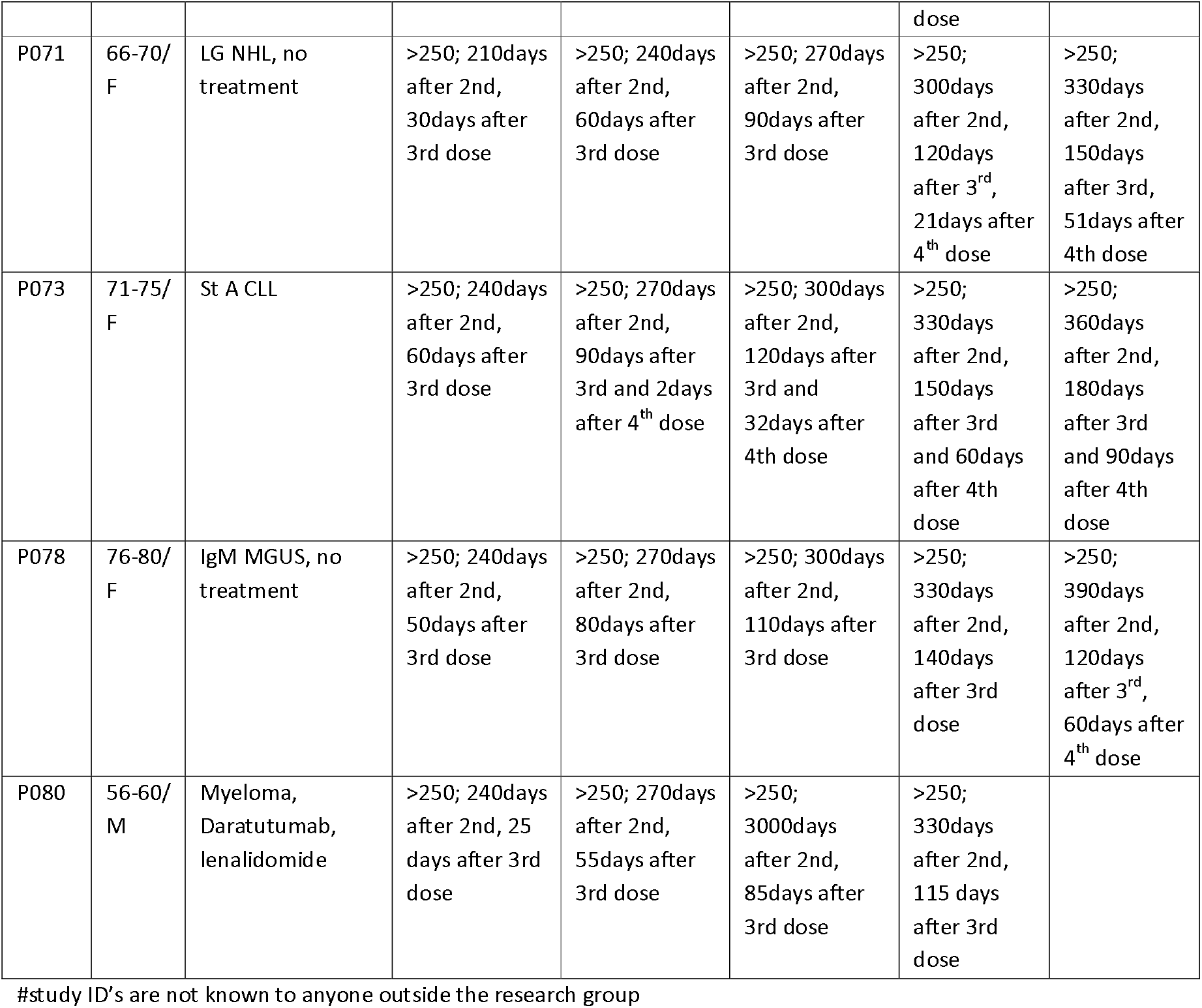
Details of study group (immunocompromised) patients with good antibody response following Covid-19 vaccination

| Study ID# | Age range (yr)/ Sex | Clinical (diagnosis; treatment) | Total Antibody to Spike protein (result; days after second dose of covid vaccine) (Positive result $\geq 0.8$ U/ml) | | | | |
| --- | --- | --- | --- | --- | --- | --- | --- |
|  |  |  | BT1 | BT2 | BT3 | BT4 | BT5 |
| P011 | 61-65/<br>M | Myeloma; VTD, daratumumab, lenalidomide | >250; 30 days after 2<sup>nd</sup> dose | ND | >250; 60 days after 2<sup>nd</sup> dose | >250; 90 days after 2<sup>nd</sup> dose | >250; 120 days after 2<sup>nd</sup> dose |
| P023 | 51-55/<br>F | Myeloma; lenalidomide | >250; 30 days after 2<sup>nd</sup> dose | ND | >250; 60 days after 2<sup>nd</sup> dose | >250; 90 days after 2<sup>nd</sup> dose | >250; 120 days after 2<sup>nd</sup> dose |
| P028 | 16-20/<br>M | ALPS; no treatment | >250; 30 days after 2<sup>nd</sup> dose | ND | >250; 60 days after 2<sup>nd</sup> dose | >250; 90 days after 2<sup>nd</sup> dose | >250; 120 days after 2<sup>nd</sup> dose |
| P032 | 31-35/<br>F | Myeloma; lenalidomide | >250; 15 days after 2<sup>nd</sup> dose | >250; 30 days after 2<sup>nd</sup> dose | >250; 60 days after 2<sup>nd</sup> dose | >250; 90 days after 2<sup>nd</sup> dose | >250; 120 days after 2<sup>nd</sup> dose |
| P038 | 71-75/<br>F | Acute Myeloid Leukaemia | >250; 15 days after 2<sup>nd</sup> dose | >250; 30 days after 2<sup>nd</sup> dose | >250; 60 days after 2<sup>nd</sup> dose | >250; 90 days after 2<sup>nd</sup> dose | >250; 120 days after 2<sup>nd</sup> dose |
| P045 | 71-75/<br>M | Myeloma, post ASCT, lenalidomide maint | >250; 150days after 2<sup>nd</sup> dose | >250; 180days after 2<sup>nd</sup> dose | >250; 210days after 2<sup>nd</sup> and 45days after 3<sup>rd</sup> dose | >250; 240days after 2<sup>nd</sup>, 75days after 3<sup>rd</sup> dose | >250; 270days after 2<sup>nd</sup>, 105 days after 3<sup>rd</sup> dose |
| P064 | 56-60/<br>M | Stage A CLL, no Treatment | >250; 180days after 2<sup>nd</sup> dose, | >250; 210days after 2<sup>nd</sup> dose, 20days after 3<sup>rd</sup> dose | >250; 240days after 2<sup>nd</sup> dose, 50days after 3<sup>rd</sup> dose | >250; 270days after 2<sup>nd</sup> dose, 80days after 3<sup>rd</sup> dose | >250; 300days after 2<sup>nd</sup>, 110days after 3<sup>rd</sup> dose |
| P066 | 66-70/<br>M | Myeloma, Daratumumab, Velcade, Dex | >250; 180days after 2<sup>nd</sup> dose | >250; 210days after 2<sup>nd</sup> dose, 30days after 3<sup>rd</sup> dose | >250; 240days after 2<sup>nd</sup> dose, 60days after 3<sup>rd</sup> dose | >250; 270days after 2<sup>nd</sup> and 90days after 3<sup>rd</sup> dose | >250; 300days after 2<sup>nd</sup> and 120 days after 3<sup>rd</sup> dose |
| P069 | 71-75/<br>M | Stage A CLL, no Treatment | >250; 180days after 2<sup>nd</sup> dose | >250; 210days after 2<sup>nd</sup> dose, 45days after 3<sup>rd</sup> dose | >250; 240days after 2<sup>nd</sup>, 75days after 3<sup>rd</sup> dose | >250; 270days after 2<sup>nd</sup>, 105days after 3<sup>rd</sup> dose | >250; 300days after 2<sup>nd</sup>, 135days after 3<sup>rd</sup> and 30days after 4<sup>th</sup> dose |
| P070 | 66-70/<br>F | Myeloma, Cardiac Amyloidosis; Daratumumab, velcade, dex | >250; 180days after 2<sup>nd</sup> dose, 30days after 3<sup>rd</sup> | >250; 210days after 2<sup>nd</sup> and 60days after 3<sup>rd</sup> dose | >250; 240days after 2<sup>nd</sup>, 90days after 3<sup>rd</sup> and 2 days after 4<sup>th</sup> dose | >250; 270days after 2<sup>nd</sup>, 120days after 3<sup>rd</sup> and 32 days after 4<sup>th</sup> dose | >250; 300days after 2<sup>nd</sup>, 150days after 3<sup>rd</sup> and 60days after 4<sup>th</sup> dose |
| P071 | 66-70/<br>F | LG NHL, no<br>treatment | >250; 210days<br>after 2nd,<br>30days after<br>3rd dose | >250; 240days<br>after 2nd,<br>60days after<br>3rd dose | >250; 270days<br>after 2nd,<br>90days after<br>3rd dose | >250;<br>300days<br>after 2nd,<br>120days<br>after 3<sup>rd</sup>,<br>21days after<br>4<sup>th</sup> dose | >250;<br>330days<br>after 2nd,<br>150days<br>after 3rd,<br>51days after<br>4th dose |
| P073 | 71-75/<br>F | St A CLL | >250; 240days<br>after 2nd,<br>60days after<br>3rd dose | >250; 270days<br>after 2nd,<br>90days after<br>3rd and 2days<br>after 4<sup>th</sup> dose | >250; 300days<br>after 2nd,<br>120days after<br>3rd and<br>32days after<br>4th dose | >250;<br>330days<br>after 2nd,<br>150days<br>after 3rd<br>and 60days<br>after 4th<br>dose | >250;<br>360days<br>after 2nd,<br>180days<br>after 3rd<br>and 90days<br>after 4th<br>dose |
| P078 | 76-80/<br>F | IgM MGUS, no<br>treatment | >250; 240days<br>after 2nd,<br>50days after<br>3rd dose | >250; 270days<br>after 2nd,<br>80days after<br>3rd dose | >250; 300days<br>after 2nd,<br>110days after<br>3rd dose | >250;<br>330days<br>after 2nd,<br>140days<br>after 3rd<br>dose | >250;<br>390days<br>after 2nd,<br>120days<br>after 3<sup>rd</sup>,<br>60days after<br>4<sup>th</sup> dose |
| P080 | 56-60/<br>M | Myeloma,<br>Daratumumab,<br>lenalidomide | >250; 240days<br>after 2nd, 25<br>days after 3rd<br>dose | >250; 270days<br>after 2nd,<br>55days after<br>3rd dose | >250;<br>3000days<br>after 2nd,<br>85days after<br>3rd dose | >250;<br>330days<br>after 2nd,<br>115 days<br>after 3rd<br>dose |  |

#study ID's are not known to anyone outside the research group

There were no reports of grade 2 or higher adverse events following vaccination. Only local AE were reported by 12 participants with no difference between the two groups. No participant suffered from Vaccine induced thrombocytopenia and thrombosis.

It is possible the inability to identify significant differences between study and control groups was due to small sample size, clinical heterogeneity, different time points of vaccination and samples taken.

No participant died or was hospitalised due to severe Covid-19 infection during the study period. This included study group participants who had no antibody response at any time point.

**Fig 1:**
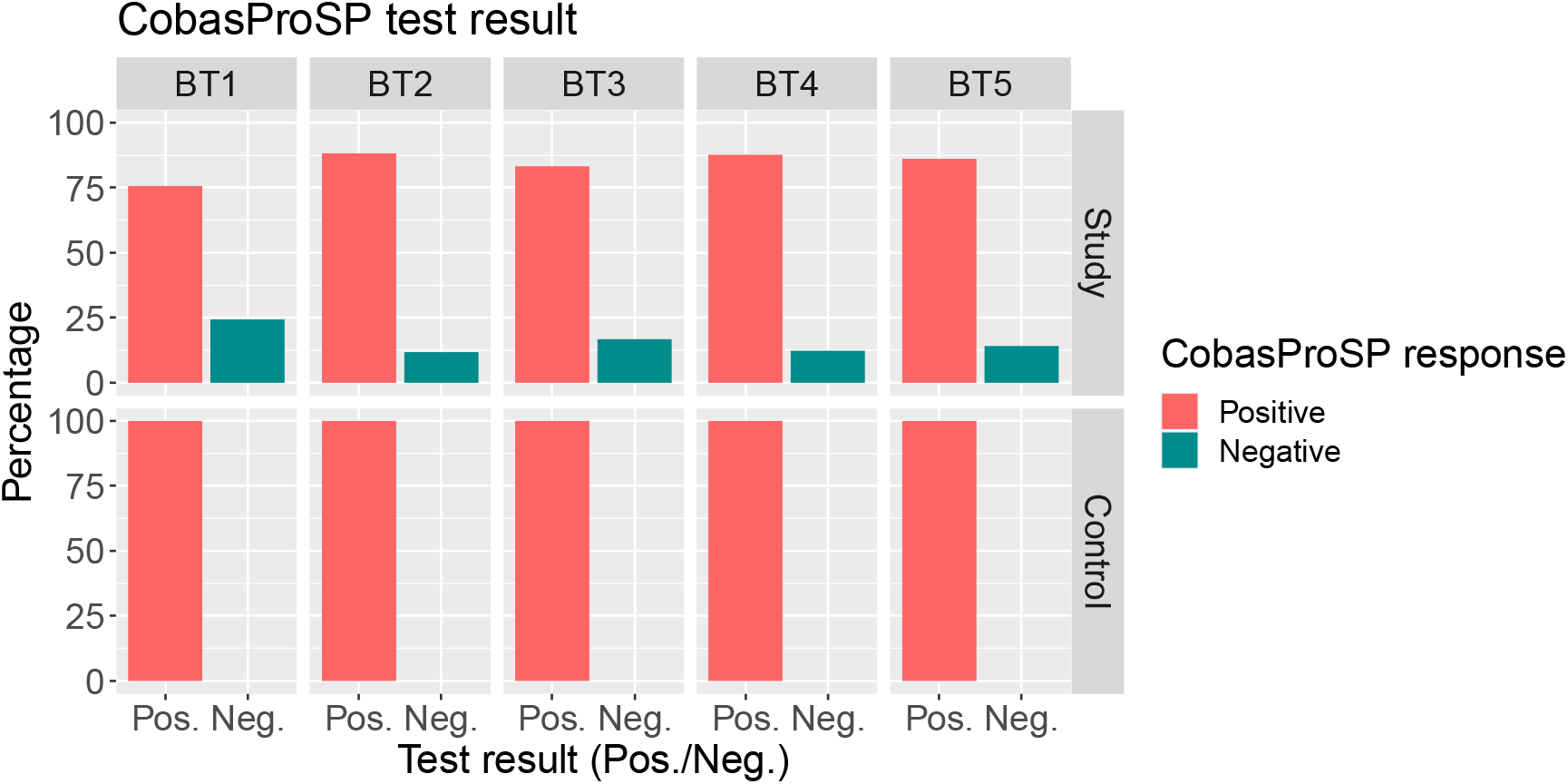
Comparison of Total Anti-SARS-Cov-2 S antibody positive and negative response between study (SG) and control group (CG)

**Fig 2:**
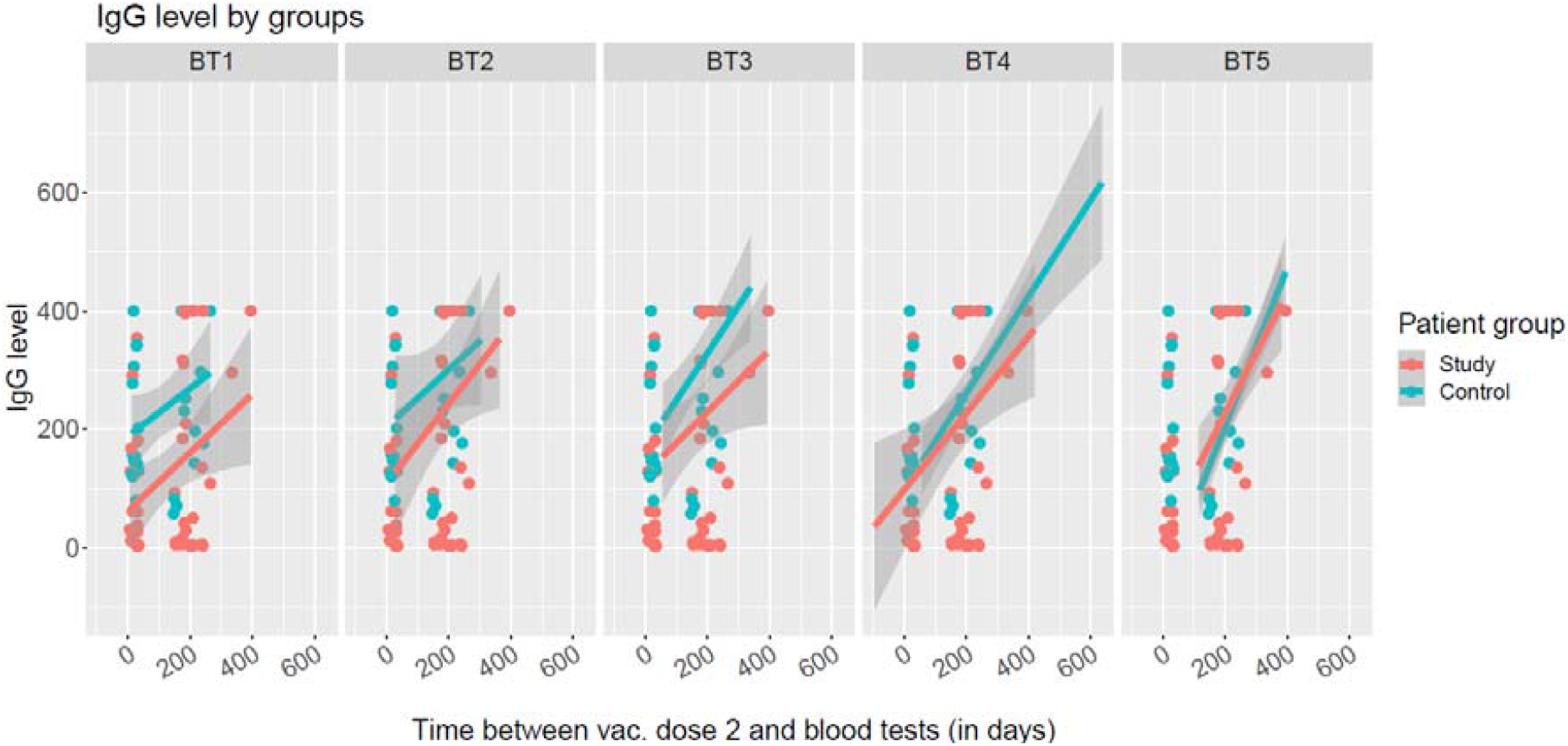
Total Anti-SARS-Cov-2 S antibody response trend from time after 2^nd^ covid vaccination

## Discussion

SARS-CoV-2 vaccines have proven highly effective in preventing severe Covid-19 but there remains considerable concern about their efficacy in haematological immunocompromised patients [3].

This prospective study evaluated immune response and safety in immunocompromised individuals with haematological diseases who had received at least 2 doses of Covid-19 vaccination and compared it with an immunocompetent control group.

When planning our study in Jan 2021, we wanted rapid confirmation of clinical efficacy and safety of these vaccines in immunocompromised haematology patients. There was uncertainty about the response to the vaccine and fear of side effects particularly after reports of vaccine induced thrombosis and thrombocytopenia [4]. We chose to measure serologic response with the assumption that this will be the best correlate for clinical efficacy. The study assessed immunological response at 30 day time intervals after the second dose of Covid-19 vaccination. Blood samples were tested for antibody response was tested up to 5 time points at 30 day intervals. All participants who were able to get at least 3 tests were included in the analysis. T cell response was checked at 1^st^ blood test. It was intended to perform T cell response assessment at 3time points however shortage of kit during the pandemic did not allow this to happen. There were significant challenges during the conduct of the study. The public health vaccination schedule changed the vaccine dose interval from 21-28 days to 10-12 weeks. To allow study recruitment the trial management group (TMG) had to amend the study to allow participation of all individuals who had received at least 2 doses of vaccine at least 30 days before their 1^st^ blood test and thereafter tested samples at 30 day intervals. Public health changes to vaccination happened again introducing 3^rd^ and 4^th^ doses for immunocompromised individuals. This resulted in some blood samples being taken after 3^rd^ and 4^th^ doses. These changes introduced considerable heterogeneity in the study.

All participants received either ChAdOx1-S (AstraZeneca) or BNT162b2 mRNA vaccine (Pfizer). No participant reported any grade II or more side effects. Grade I side effects between the two groups were comparable.

Several investigators have studied humoral and T cell response following one or two doses of Covid-19 vaccine in people with haematological malignancies. Overall seropositivity rate reported in terms of anti-SARS-Cov-2 S antibody was 62-66% after 2 doses of Covid-19 vaccine. The neutralizing antibody response rate was 57-60% though this declined with emergence of new covid-19 variants of concern. The T cell cellular response rate reported in these studies varied between 40-75% [5]. This was lower than the vaccine efficacy reported in immunocompetent individuals of 74-95% [6]

Vaccine induced antibodies against the spike domain have potent SARS-Cov-2 neutralizing activity [7]. Some researchers have proposed higher cut-off values for serologic response to better correlate with neutralising antibody response. This however varied between different assays and was also affected by the emergence of new covid variants [6, 8].

Our study confirmed that 13% of immunocompromised individuals had no detectable Anti-SARS – Cov-2 S antibody response at any time point. Three (7%) of study group participants had no response even after additional booster doses of Covid-19 vaccine.

We found a non-significant difference in T cell and total Anti-SARS-CoV-2 S antibody response between study and control group patients.

Different factors including age, underlying haematological condition, immunosuppressive treatment (and its timing), vaccine type, number of doses, timing of blood test and natural infection would have affected our results. Due to this heterogeneity, definite response determinants are difficult to identify. We compared immunocompromised subgroups with poor or good antibody response. No specific haematological diagnosis, chemotherapy or immunotherapy was identified as determinant of response.

Similar findings have been reported by several investigators. In a meta-analysis of more than 7000 patients with haematologic malignancies statistical heterogeneity was substantial in more than 70%. Studies were clinically heterogeneous because of different haematologic malignancies, lack of standardised platforms and variable follow-up periods. Patients with haematologic malignancies are reported to have a lower serologic response following 2 doses of Covid-19 vaccination [5]. This response is improved following 3^rd^ and 4^th^ doses of covid vaccine [9]. What is unclear however, is whether a positive total Anti-SARS-CoV-2 S antibody correlates with vaccine efficacy. None of the studies have reported clinical efficacy in terms of incidence of severe Covid-19 disease or mortality between immunocompromised individuals with and without antibody response. In our cohort no study group participant suffered from severe disease or mortality due to covid. While this could be due to small sample size or strict isolation followed by immunocompromised individuals, the possibility remains that T-cell immunity provides considerable protection from severe disease and Covid-19 mortality. With emergence of new Covid-19 variants the serologic response in terms of neutralising antibodies declined. T-cell responders however had a better protection against Covid-19 variants of concern [10].

Covid-19 anti-spike antibodies can be considered an indicator of immune response but are not a reliable correlate of clinical protection. In the majority of studies, including our own, the outcome of interest was seropositivity, which does not equate to seroprotection. There is lack of standardised thresholds and variability across different commercial assays and platforms. The neutralising antibody response is dependent on the Covid-19 variant tested against. To date there is no reliable correlate of protection that allows definite deduction of clinical efficacy from immune response generated either in the immunosuppressed or the general population [6]. Further studies should focus on identifying the correlation between serological response, T cell response, neutralisation and clinical protection from symptomatic and severe Covid-19 infection. This would enable development of standardised assays and cut-off which would correlate with clinical protection.

In conclusion our study found non-significant difference in T cell and total Anti-SARS-CoV-2 S antibody response between immunocompromised patients with haematological diseases when compared with immunocompetent adults. Covid-19 vaccination was well tolerated without major side effects in both groups. We did not identify any patient-specific factor (age, gender), specific haematological condition or treatment as determinant of response. Different vaccination times, doses, timing of blood test and natural infection with Covid-19 added considerable heterogeneity in the study. No patient suffered from severe Covid-19 infection or died due to Covid-19 during the study.

## Data Availability

All data is available upon request. All information governance regulations were followed

## Acknowledgement

This study is funded by the University Hospitals of North Midlands Research Charitable Fund. The study was sponsored by the Research and Innovation Directorate at University Hospitals of North Midlands. The investigators are grateful to Dr Simon Lea, Dr Keira Watts, Ms Karen Sneade, Ms Zuzana Tothova and Mr Jack Tilstone for their support and assistance.

